# Clinical, imaging, serological, and histopathological features of pulmonary post-acute sequelae after mild COVID-19 (PASC)

**DOI:** 10.1101/2022.11.29.22282913

**Authors:** D Gagiannis, C Hackenbroch, A Czech, A Lindner, N Maag, W Bloch, F Zech, F Kirchhoff, S Djudjaj, S von Stillfried, R Bülow, P Boor, K Steinestel

## Abstract

**Background:** A significant proportion of patients experience prolonged pulmonary, cardiocirculatory or neuropsychiatric symptoms after Coronavirus disease 2019 (COVID-19), termed post-acute sequelae of COVID (PASC). Lung manifestations of PASC include cough, dyspnea on exertion and persistent radiologic abnormalities and have been linked to viral persistence, ongoing inflammation and immune dysregulation. So far, there is limited data on lung histopathology and tissue-based immune cell subtyping in PASC.

**Methods:** 51 unvaccinated patients (median age, 40 years; 43% female) with a median of 17 weeks (range, 2-55 weeks) after mild SARS-CoV-2 infection (without hospitalization) underwent full clinical evaluation including high-resolution computed tomography (HR-CT) and transbronchial biopsy. We used RT-PCR/FISH and immunohistochemistry (nucleocapsid/spike/CD3/CD4/CD8) for residual SARS-CoV-2 detection and T lymphocyte subtyping, respectively. We assessed interstitial fibrosis and macrophage profiles by transmission electron microscopy (TEM) and immunofluorescence multiplex staining, while cytokine profiling in bronchoalveolar lavage (BAL) fluid was performed by legendplex immunoassay.

**Results:** Dyspnea on exertion was the leading symptom of pulmonary PASC in our cohort. In 16% and 42.9% of patients, FEV1 and MEF50 were ≤ 80% and 35.3% showed low attenuation volume (LAV) in >5% of lung area, in line with airflow obstruction. There was a significant correlation between oxygen pulse and time since COVID (p=0.009). Histopathologically, PASC manifested as organizing pneumonia (OP), fibrinous alveolitis and increased CD4+ T cell infiltrate predominantly around airways (bronchiolitis), while the residual virus components were detectable in only a single PASC patient (2%). T cell infiltrates around small airways were inversely correlated with time since COVID, however, this trend failed to reach statistical significance. We identified discrete interstitial fibrosis and a pro-fibrotic macrophage subtype (CD68/CD163/S100A9) as well as significantly elevated interleukin 1β in BAL fluid from PASC patients (p=0.01), but H-scores for fibrotic macrophage population did not correlate with severity of clinical symptoms or T cell infiltration.

**Interpretation:** We show decreased FEV1/MEF50 and increased LAV in line with obstructive lung disease due to CD4+ T cell-predominant bronchiolitis as well as evidence of pro-fibrotic signaling in a subset of unvaccinated PASC patients. Since our results point towards self-limiting inflammation of small airways without detectable viral reservoirs, it remains unclear whether pulmonary symptoms in PASC are SARS-CoV-2-specific or represent a general response to viral infection. Still, evidence of pro-fibrotic signaling should warrant clincal follow-up and further research into possible long-time fibrotic remodeling in PASC patients.

**Key points:**

- Dyspnea on exertion is the leading clinical manifestation of PASC in the lung
- a minority of pts have significantly impaired lung function (FVC/TLC≤80% or DLCO≤70%) in spiroergometry and/or radiologic abnormalities, oxygen pulse seems to normalize over time
  - 16% and 42.9% of pts have FEV1 and MEF50≤80% and 35.3% have LAV>5% of lung area, in line with airflow obstruction due to bronchiolitis
- Residual virus was not detectable in the lung tissue of all but one PASC patient (2%)
- Histologically, PASC may manifest as T cell-mediated bronchiolitis, OP and fibrinous alveolitis
- There is evidence of fibrotic remodeling (ultrastructural interstitial fibrosis, pro-fibrotic macrophage subpopulation, pro-fibrotic cytokine IL-1β in BAL) but this did not correlate with the degree of T cell infiltrate/bronchiolitis

## INTRODUCTION

Following SARS-CoV-2 infection, a subgroup of patients develop lasting symptoms, termed post-acute sequelae of COVID-19 (PASC). The burden of PASC is associated with the severity of the initial disease and lies between 4% and 15% at 6 to 8 months among non-hospitalized patients[1-3]. Lung manifestations of PASC include shortness of breath, chest pain and imaging abnormalities such as ground glass opacities (GGO) and subpleural bands [4]. Concerning pulmonary function tests, a systematic review and meta-analysis reported altered diffusion lung capacity for carbon monoxide (DLCO), and restrictive and obstructive patterns in PASC patients, but the test results did not correlate with the subjective severity of symptoms in another study [5,6]. Concerning histopathology, one study did not find distinct post-acute histopathologic changes in lung tissue of elective lung resection specimens from patients after mild and moderate COVID-19, however, these patients did not show PASC symptoms [7]. Another case study reported extensive interstitial fibrosis in a lung explant from a 62-year-old man 12 weeks after recovering from mild COVID-19 [8]. While it is clear that the long-term outcome in patients after severe COVID-19 (SARS-CoV-2-associated acute respiratory distress syndrome, ARDS, and/or who underwent mechanical ventilation) might be explained by widespread lung damage, the pathophysiology of pulmonary sequelae in mild cases of COVID-19 is unclear. It has been proposed that impaired lung function might be associated with dysregulated respiratory CD8+ T cell responses or damage to the pulmonary vasculature after acute COVID-19 [9,10]. Furthermore, presence of circulating antinuclear autoantibodies (ANA) have been shown to predict PASC in COVID-19 survivors [11]. This is of interest given the association between the presence of autoantibodies (ANA/ENA) and the severe course of acute COVID-19, which we have previously described [12]. Another hypothesis for the pathogenesis of PASC is based on a possible persistence of SARS-CoV-2 or viral particles/mRNA in tissue, driving chronic inflammation [13]. Verification of each of these hypotheses is hampered by the lack of data on tissue samples from PASC patients in most published studies. The present multidisciplinary study aimed to characterize clinical, imaging, serological and histopathological features in a consecutive cohort of PASC patients from our postCOVID outpatient clinic.

## METHODS

### Patients and diagnostic work-up

We included n=66 patients with a history of RT-PCR-confirmed SARS-CoV-2 infection and persistent pulmonary symptoms in the present study. All patients were unvaccinated at the time of SARS-CoV-2 infection (before vaccine roll-out) and had a mild clinical course (without hospitalization). The baseline diagnostic workup included clinical history including current and previous medication and allergies, physical examination and spiroergometry. ANA/ANCA/ENA screening was performed as previously described [12]. After multidisciplinary discussion, bronchoscopy with transbronchial biopsy for the assessment of possible PASC-ILD was performed in n=51 patients who formed the definite cohort. Patients have given written informed consent to routine diagnostic procedures (serology, bronchoscopy, imaging) as well as to the scientific use of data and tissue samples in the present study. This project was approved by the local ethics committee of the University of Ulm (ref. no. 129-20) and conducted in accordance with the Declaration of Helsinki.

### Imaging

Imaging of the lung was performed on a 3rd generation DECT Scanner (Siemens Somatom Force, Siemens Healthineers, Forchheim, Germany). A non-contrast multislice spiral CT in fulldose technique (Qual. Ref. kV: Sn 100, Qual. Ref. mAs: 600, ref. CTDI 1.06 mGy) with a collimation of 192 × 0.6 mm and using a tin prefilter at 100 resp. 150 kV was obtained. Rotation time 0.25 sec, scan time 1.25 sec. Examination was performed in inspiration. Reconstruction in bone, soft tissue and lung window. The subsequent automated segmentation and evaluation of the lung volumes as well as the determination of overinflated or fibrotically altered/attenuated lung areas was carried out with the manufacturer-specific evaluation software “syngo.via”, version VB 40 (Siemens Healthineers, Forchheim, Germany) using the software “CT Pulmo 3D”, with a reconstruction set of the acquired CT scan with the following specifications: Slice thickness 0.75 mm, increment 0.5 mm, reconstruction with a medium soft kernel Br 40, iterative reconstruction: Admire stage 3. A threshold value of minimum -950 HU (low attenuation value, LAV) and maximum -200 HU (high attenuation value, HAV) was defined as the limits of regularly ventilated lung areas. Values below LAV were considered overinflated areas, values above HAV were considered attenuated lung areas.

### Histology and SARS-CoV-2 detection

Lung tissue specimens were obtained as transbronchial biopsies and stained with haematoxylin-eosin (HE), Elastica-van-Gieson (EvG) and Masson-Goldner (MG). Slides were reviewed with special emphasis on possible post-viral change (alveolitis, peribronchiolitis, airway smooth muscle hypertrophy, goblet cell hyperplasia, airway squamous epithelial metaplasia, and fibrosis)[14]. We performed immunohistochemistry for Spike and Nucleocapsid proteins of SARS-CoV-2 using the following antibodies: anti-SARS-CoV-2 spike antibody, clone 1A9, mouse monoclonal, 1:400 (GeneTex, Irvine, CA, USA); anti-SARS-CoV-2 nucleocapsid antibody, rabbit monoclonal, 1:20000 (SinoBiological, Bejing, China). Autopsy-derived tissue samples from deceased COVID-19 patients (**Fig. 2 B**, upper panel) served as positive controls [15]. For SARS-Cov-2 fluorescence in situ hybridization (FISH), paraformaldehyde-fixed, paraffin-embedded 1µm sections of transbronchial biopsies (TBBs) and lung autopsy material (positive controls) were deparaffinized followed by dehydration with 100% ethanol. FISH was performed with the RNAscope® Multiplex Fluorescent Reagent Kit v2 assay (Advanced Cell Diagnostics, Inc.) according to the manufacturer’s instruction. Briefly, a heat-induced target retrieval step followed by protease was performed. Afterwards sections were incubated with the following RNAscope® Probe -V-nCoV2019-S (#848561-C1), -V-nCoV2019-S-sense (#845701-C1), -Hs-ACE2-C2 (#848151-C2) and -Hs-TMPRSS2-C2 (#470341-C2). After the amplifier steps, the fluorophores OpalTM 570 and 650 (PerkinElmer Life and Analytical Sciences) were applied to the tissues incubated with C1 and C2 probes, respectively. Finally, nuclei were stained with DAPI and the slides were mounted with ProLongTM Gold antifade reagent (Invitrogen). Sections were analyzed with Zeiss Axio Imager 2 and image analysis software (ZEN 3.0 blue edition). SARS-CoV-2 RNA was extracted using Maxwell® 16 FFPE Plus Tissue LEV DNA Purification KIT (Promega) on Maxwell® 16 IVD Instrument (Promega). Using TaqMan® 2019-nCoV kit (ThermoFisher), the detection of SARS-CoV-2 E-gene was performed according to the manufacturer’s instructions.

**Figure 1.**
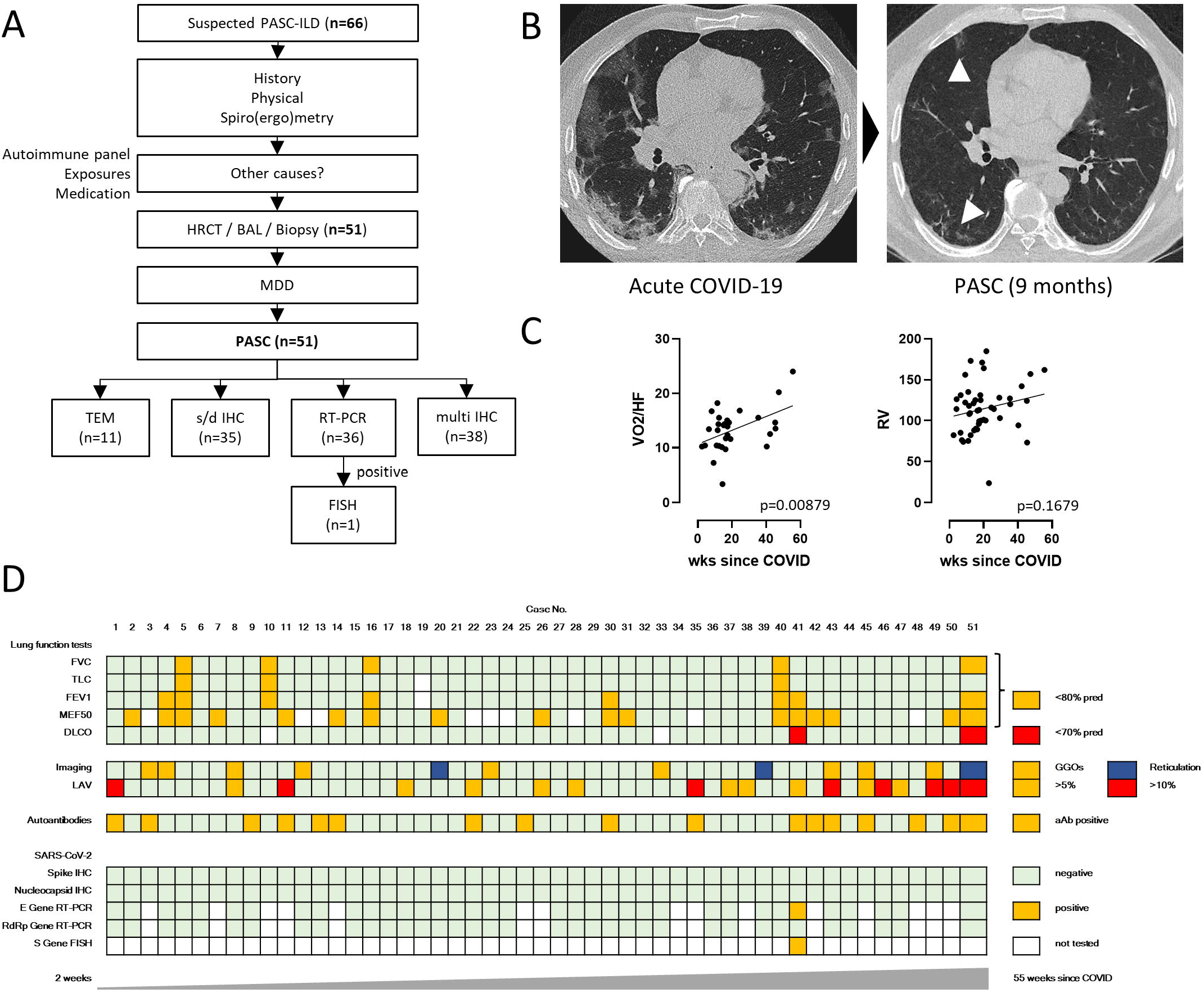
Characterization and diagnostic work-up of PASC patients. **A**, Flow diagram for diagnostic work-up of suspected PASC-ILD patients. Lung biopsies from 51 PASC patients were finally included in histopathological assessment. **B,** representative imaging of acute COVID-19 (left) and PASC (right) with discrete reticulation and ground glass opacities (arrowheads, 9 months post-infection). **C,** correlation between oxygen pulse (left, p<0.01) and residual volume (right, p=0.1679) and time since COVID-19, indicating restoration of lung function over time. **D,** heatmap stratified according to time post infection (2 wks to 55 wks) displaying the results from lung function tests, imaging, autoantibody screening and tissue-based SARS-CoV-2 detection. FISH analysis was only performed in the single patient with positive SARS-CoV-2 RT-PCR (case #41).

**Figure 2.**
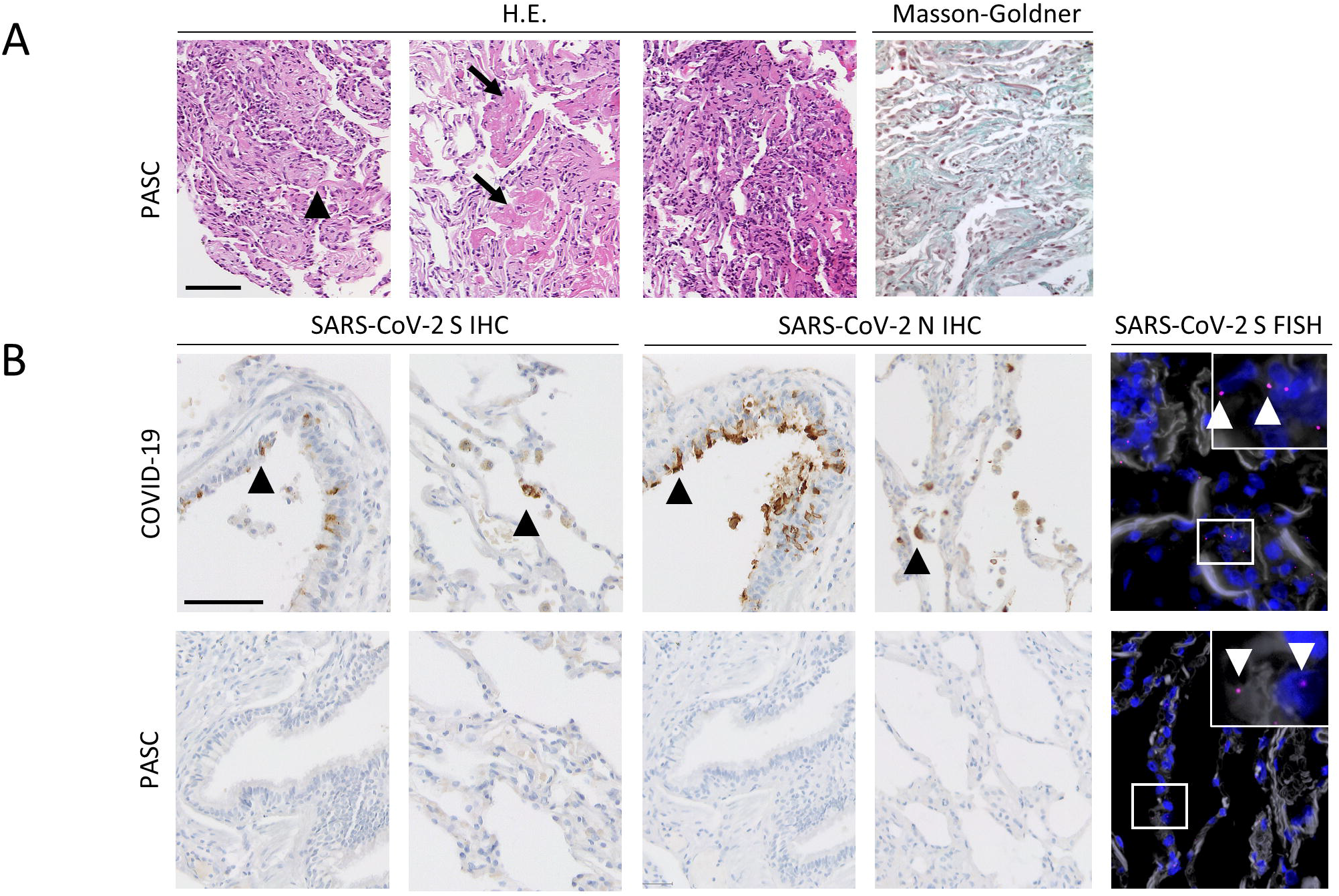
Histology and tissue-based SARS-CoV-2 detection. **A**, Representative microphotographs of H.E.-stained lung tissue samples from PASC patients showing organizing pneumonia (arrowhead), alveolar fibrinous exudate (arrows) and interstitial lymphocytosis (right image). Masson-Goldner staining showing minimal interstitial fibrosis. **B,** Representative microphotographs of immunohistochemistry for SARS-CoV-2 Spike (S) and Nucleocapsid (N) proteins in airways and lung alveoli in acute COVID-19 (upper panel) and PASC (lower panel). Both IHC stainings were negative in all PASC cases. SARS-CoV-2 S FISH showed sparse signals in alveolar epithelium the single patient with positive SARS-CoV-2 RT-PCR (case #41). *Scale bar, 100µm*.

### Electron microscopy

Lung tissue was immersion-fixed with 4% paraformaldehyde in 0.1M PBS, pH 7.4. After several washing steps in 0.1M PBS, tissue was osmicated with 1% OsO4 in 0.1 M cacodylate and dehydrated in increasing ethanol concentrations. Epon infiltration and flat embedding were performed following standard procedures. Methylene blue was used to stain semithin sections of 0.5 µm. Seventy to ninety-nanometer-thick sections were cut with an Ultracut UCT ultramicrotome (Fa. Reichert) and stained with 1% aqueous uranylic acetate and lead citrate. Samples were studied with a Zeiss EM 109 electron microscope (Fa. Zeiss) coupled to a TRS USB (2048×2048, v.596.0/466.0) camera system with ImageSP ver.1.2.6.11 (x64) software.

### Staining Macrophage Panel

Slides underwent antigen retrieval in citrate buffer (EnVision FLEX TARGET RETRIEVAL SOLUTION LOW pH, from Agilent: K8005) using the pT-Link module (Agilent, Santa Clara, USA). After fixation in 4% formalin for 10 min, slides were washed and blocking was performed with H2O2 (DAKO REAL PEROXIDASE-BLOCKING SOLUTION, Agilent, Santa Clara, USA: S2023) followed by 30 min incubation with antibody diluent (DAKO REAL ANTIBODY DILUENT, Agilent, Santa Clara, USA: S2022). Immunofluorescence multiplex staining was performed with Opal 7-Color Manual IHC Kit (AKOYA Biosciences, Menlo-Park, USA: NEL811001KT). The slides were incubated for 1 hour with primary antibodies: CD68 (Agilent, Santa Clara, USA: M0876), CD163 (Cell Marque: 163M-17), S100A9 (Abcam: ab63818) and CD16 (Santa Cruz Biotechnology, Dallas, USA: DJ130c), followed by incubation with EnVision FLEX HRP (Agilent, Santa Clara: DM802) and visualized with Opal 690 TSA Plus, using Opal 650 TSA Plus, Opal 620 TSA Plus, and Opal 570 TSA Plus, respectively (all from AKOYA Biosciences, Marlborough, USA). The nuclei were counterstained using Spectral DAPI (AKOYA Biosciences, Marlborough, USA).

### Macrophage phenotyping

We scanned twelve regions of interest of lung tissue to multispectral images (MSI) per sample using the 40x objective, corresponding to a tissue area of 334×250 µm^2^ each. Scanning was performed using the VECTRA automated quantitative pathology imaging system (Perkin Elmer, Waltham, USA). After deploying automated cell detection using the InForm Software (Perkin Elmer, Waltham, USA), we trained an in-built cell phenotyping algorithm to detect CD16+, CD16/CD163+, CD68+ and CD68+/CD163+ cells, as well as a final cell type containing all other cells. Additionally, we assessed S100A9-Expression in the detected and classified cells automatically in four bins (0-3) and calculated and H-Score of S100A9-Expression. After training all samples were analyzed in one batch using the finale algorithm. Measurement outputs of the InForm Software were analyzed in R version 4.0.3 using the phenoptr (v0.3.2) and phenoptrReports (v0.3.3) packages.

### BAL analysis and legendplex immunoassay

The Legendplex ELISA was performed according to the manufacturer’s instructions. In brief, 25 μl of the collected BAL was centrifuged (4000 rpm, 5 min) and incubated for 2 h at room temperature with antibody-coated beads, followed by washing and incubation with the detection antibodies. After incubation with the staining reagent, the beads were analyzed in a high-throughput sampler via flow cytometry (FACSCanto II, BD Biosciences). Absolute quantification was performed using a standard and the Bio-legend Legendplex v8.0 software.

### Statistical Methods

Descriptive statistical methods were used to summarize the data. Medians and interquartile ranges were used to announce results. Absolute numbers and percentages were employed to represent categorial variables. Student’s t-test/ANOVA with multiple comparison post-test was used for the comparison of continuous variables, while Chi-Square-Test/Fisher’s test was used for categorial variables. All statistical analyses were conducted using GraphPad PRISM 9.4.1 (GraphPad Software Inc., San Diego, CA, USA). A p-value <0.05 was regarded as statistically significant.

### Illustrations

Parts of Fig. 3 and 4 were drawn by using pictures from Servier Medical Art. Servier Medical Art by Servier is licensed under a Creative Commons Attribution 3.0 Unported License (https://creativecommons.org/licenses/by/3.0/).

**Figure 3.**
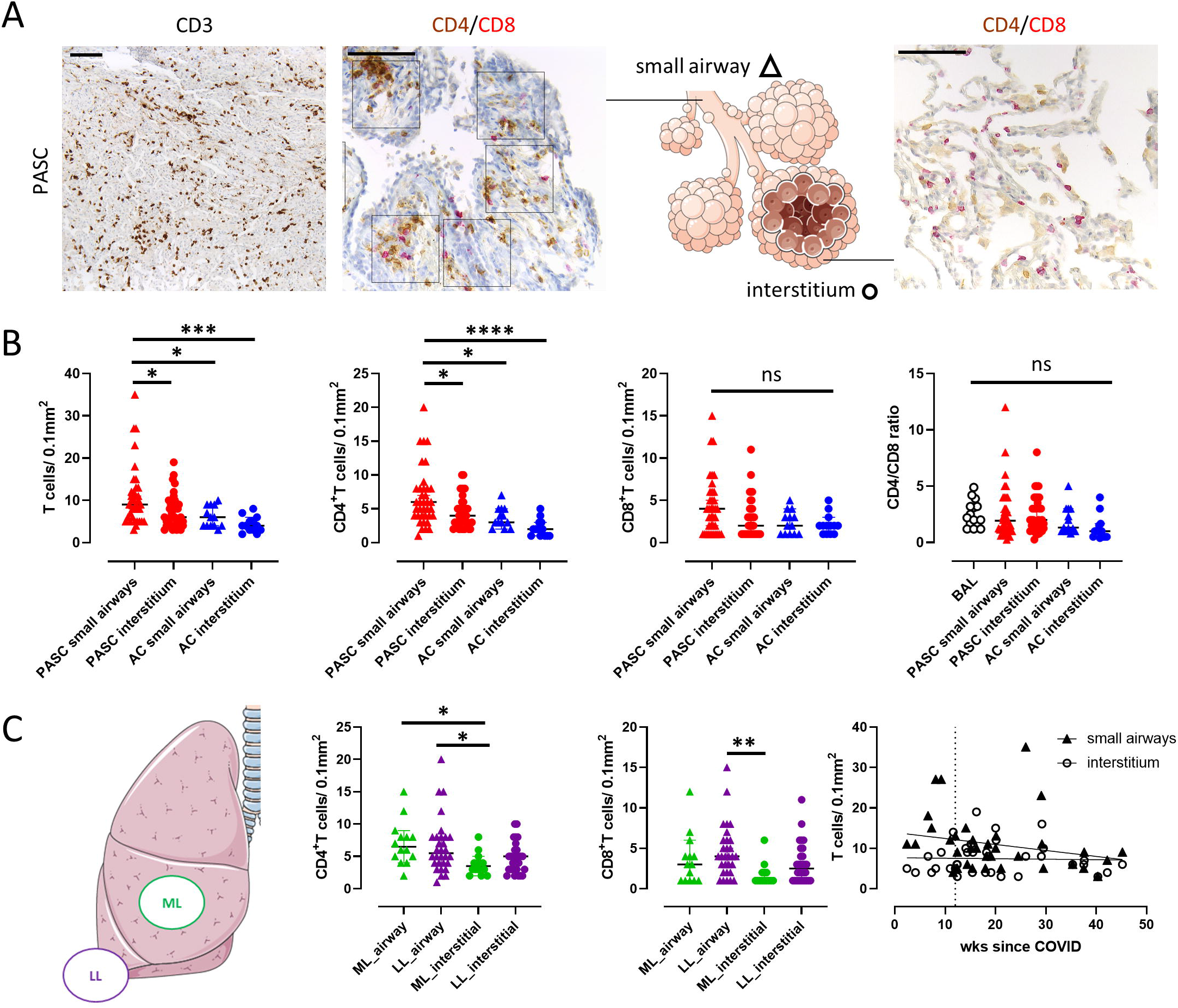
Spatial T lymphocyte subtyping in PASC patients. **A**, representative microphotographs of immunohistochemistry for pan-T-cell-marker (CD3) and CD4/CD8 double-staining in small airways (triangle) and lung alveoli (circle). **B,** statistical analyses showed significantly elevated numbers of CD3+ T-cells and CD4+ T-helper cells, but not CD8+ cytotoxic T-cells, in small airways compared to interstitium in PASC patients as well as compared to both small airways and interstitium of pre-pandemic autopsy controls. CD4/CD8 ratio was not significantly different compared to both PASC BAL fluid and pre-pandemic autopsy controls. **C,** distribution of CD4+/CD8+ T-cells between different biopsy sites (middle lobe/lower lobe) and correlation between T-cell infiltration and time since COVID-19. There was a decrease in the number of airway T-cells over time.

**Figure 4.**
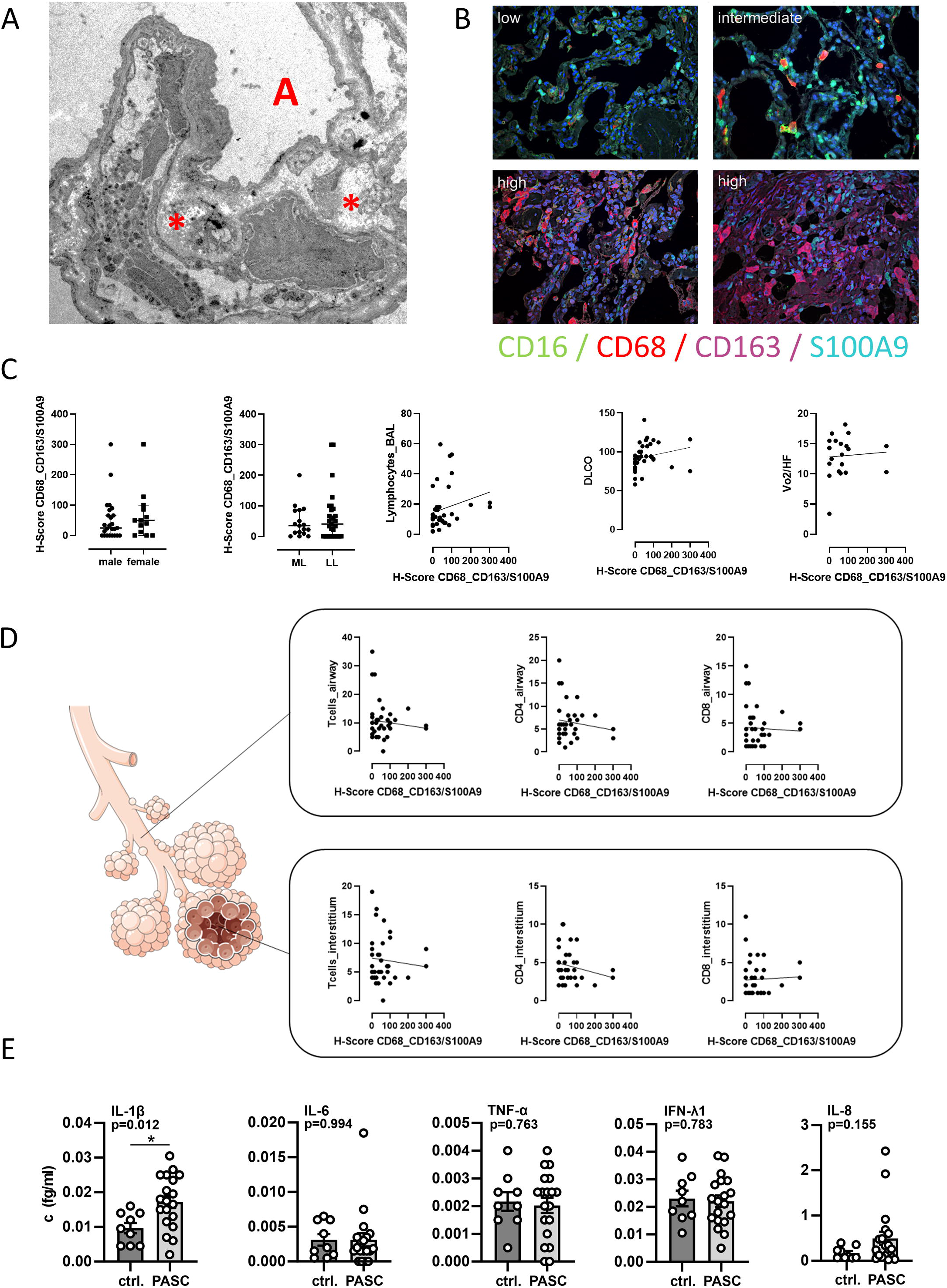
Assessment of interstitial fibrosis and quantification of macrophage subpopulations in PASC patients. **A**, TEM shows interstitial deposition of collagen fibrils. **B,** representative microphotographs of immunofluorescence multiplex staining of lung tissue samples. *Scale bar, 100µm*. **C,** H-scores for pro-fibrotic macrophage phenotype (CD68/CD163/S100A9) were not significantly different between men and women or between biopsy site (middle lobe vs. lower lobe). There was no significant correlation between H-scores for pro-fibrotic macrophage phenotype and between lymphocyte count in BAL, diffusion capacity for carbon monoxide or oxygen pulse. **D,** no significant correlation between H-scores for pro-fibrotic macrophage phenotype and CD3+ T-cells/ CD4+ T-helper cells/ CD8+ cytotoxic T-cells in both small airways and interstitium. **E,** legendplex immunoassay from BAL fluid shows elevated levels of IL-1β (p<0.05) and IL-8 compared to healthy controls, although the latter result did not turn out statistically significant.

## RESULTS

### Diagnostic workup, clinical data and imaging results

The diagnostic work-up of suspected PASC-ILD patients who presented to our outpatient clinic at the Bundeswehrkrankenhaus (army hospital) Ulm in 11/2020-04/2021 is summarized in **Fig. 1 A. Tab. 1** summarizes clinical and serological data of the PASC cohort who underwent transbronchial biopsy (n=51) and the independent cohort of BAL samples for legendplex immunoassay (n=18). All patients were unvaccinated and had not been hospitalized for COVID-19, 25/51 patients were soldiers (37.9%). Results from imaging were heterogeneous with ground glass opacities (GGO) in 9/51 and reticulations in 3/51 pts (17.6% and 5.9%), respectively (**Fig. 1 B**). Areas with low attenuation volume (LAV, below -950 HU) comprised more than 5% of the lungs in 18/51 pts (35.3%), of which in 8/51 pts (15.7%) LAV areas comprised more than 10% of the lungs. The most frequent pathological result from pulmonary function test were impaired MEF50 and FEV1 (≤ 80%) in 42.9 and 16% of patients while median MEF50 and FEV1 throughout the cohort were 88 (IQR, 66-99) and 95.5 (IQR, 85-102), respectively. When correlating the results from lung function tests (**Tab. 1**) with clinical, serological and imaging data, there was a significant association between time since infection and oxygen pulse, but not residual volume (p<0.01 and p=0.1679, respectively) (**Fig. 1 C**). All other characteristics showed no significant association. Autoantibodies (ANA titer ≥ 1:320 and/or positive immunoblot for Scl-70, PCNA, PM-Scl, dsDNA, SS-B, Histone) were detected in 17/51 pts (33.3%) but did not correlate with severity of symptoms or specific imaging (GGO, reticulation) or histopathological characteristics (OP, inflammatory infiltrate, evidence of fibrosis). **Fig. 1 D** summarizes results from lung function tests, imaging, serology, and tissue-based SARS-CoV-2 detection (see below), stratified according to time since acute COVID-19.

**Table 1.**
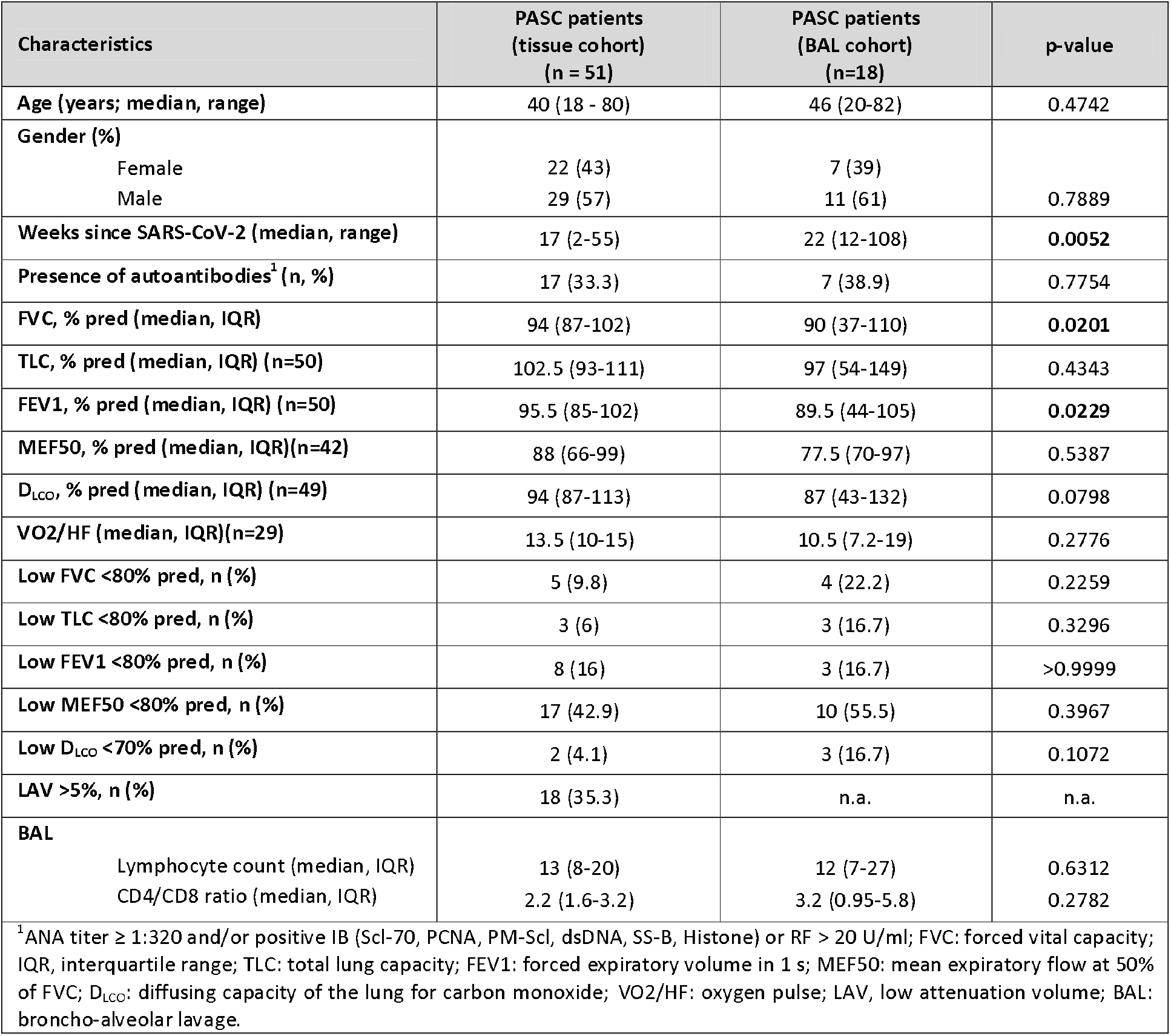
Baseline clinical characteristics and assessment of lung function in PASC patients.

### Histopathologic findings and tissue-based SARS-CoV-2 detection

Histopathologic evaluation of transbronchial biopsies from PASC patients and lung tissue samples from pre-pandemic autopsy controls (n=15, median age 56, 33% female) showed increased peribronchiolar and interstitial lymphocytosis (see below for quantification and statistical evaluation) and alveolar fibrin deposition in the PASC cohort (**Fig. 2 A** and **Tab. 2**). Organizing pneumonia was present in 2 PASC patients (3.9%) but not in autopsy controls (p>0.999). Centrilobular lymphoid follicles were observed in 47% of PASC samples and 40% of autopsy controls (p=0.558), while interstitial fibrosis (as assessed by Masson-Goldner staining and light microscopy) was present in 27% of PASC patients and 13% of pre-pandemic autopsy controls (p=0.3262). Immunohistochemistry for N and S proteins as well as RT-PCR for the RdRp gene of SARS-CoV-2 was negative in all tissue samples but one case was positive for E gene in RT-PCR (**Fig. 1 D**) and confirmed by SARS-CoV-2 S FISH analysis of the alveolar epithelium (**Fig. 2 B**).

**Table 2.** Lung histology and spatial T lymphocyte subtyping in post-COVID patients and pre-pandemic autopsy controls.

| Characteristics | PASC<br>(n = 51) | Autopsy controls<br>(n=15) | p-value |
| --- | --- | --- | --- |
| Age (years; median, range) | 40 (18 - 80) | 56 (25-82) | **0.0006** |
| Gender |  |  |  |
| Female | 22 (43%) | 5 (33%) | 0.5627 |
| Male | 29 (57%) | 10 (67%) |  |
| Organizing pneumonia |  |  |  |
| present | 2 (4%) | 0 | >0.999 |
| absent | 49 (96%) | 15 (100%) |  |
| Lymphoid follicles |  |  |  |
| present | 24 (47%) | 6 (40%) | 0.5580 |
| absent | 27 (53%) | 9 (60%) |  |
| Interstitial fibrosis |  |  |  |
| Light microscopy, n (%) | 14/51 (27%) | 2 (13%) | 0.3262 |
| TEM, n (%) | 11/11 (100%) | n.a. |  |
| Small airways (bronchi/bronchioles) (n=35) |  |  |  |
| CD4+ cells/0.1 mm<sup>2</sup> (IQR) | 6.7 (4-8) | 3.7 (3-5) | **0.0154** |
| CD8+ cells/0.1 mm<sup>2</sup> (IQR) | 4.2 (1-6) | 2.4 (1-4) | 0.1327 |
| CD4/CD8 ratio (IQR) | 2.6 (1.2-12) | 2 (1-5) | 0.8487 |
| Interstitial (alveolar septae) (n=35) |  |  |  |
| CD4+ cells/0.1 mm<sup>2</sup> (IQR) | 4.6 (3-5.5) | 2.2 (1-3) | 0.067 |
| CD8+ cells/0.1 mm<sup>2</sup> (IQR) | 2.8 (1-4) | 2.2 (1-3) | 0.8794 |
| CD4/CD8 ratio (IQR) | 2.4 (1.3-8) | 1.3 (1-2) | 0.2626 |

**Table 3.** Macrophage panel staining and automated quantitative imaging.

| Characteristics | PASC<br>(n = 38) |
| --- | --- |
| Age (years; median, range) | 39.5 (18 - 82) |
| Gender |  |
| Female | 23 (44%) |
| Male | 29 (56%) |
| Cells/mm<sup>2</sup> (median, IQR) |  |
| CD 68+ | 26 (14-46) |
| CD 16+ | 116 (17-539) |
| CD 16+/CD 163+ | 151 (72-258) |
| CD 68+/CD 163+ | 6 (1-77) |
| CD163+ Macrophages/all Macrophages (median, IQR) | 0.32 (0.15-0.79) |
| H-scores |  |
| S100A9+ (other cells) | 6 (2-22) |
| CD 68+/S100A9+ | 2 (0-9) |
| CD 16+/S100A9+ | 39 (13-61) |
| CD 16+/CD 163+/S100A9+ | 221 (123-252) |
| CD 68+/CD 163+/S100A9+ | 34 (0-85) |

### Spatial T lymphocyte subtyping in PASC patients and autopsy controls

The inflammatory infiltrates around small airways and in alveolar septa consisted of CD3+ T cells, with a predominance of CD4+ over CD8+ T cells (**Fig. 3 A**). CD4+ T cells around small airways were significantly increased compared to the interstitium in PASC patients (p=0.0205) as well as compared to both airways and interstitium of autopsy controls (p=0.0154 and p<0.0001, respectively) (**Fig. 3 B** and **Tab. 2**). There was no such difference in the distribution of CD8+ T cells between airways and interstitium as well as between PASC patients and autopsy controls (**Fig. 3 B** and **Tab. 2**). The CD4/CD8 ratio was not significantly different between BAL fluid and tissue samples (small airways and interstitium) from PASC patients or autopsy controls. When analyzing the spatial distribution of the inflammatory infiltrate in PASC patients, there was consistently more inflammation around the airways compared to the interstitium, but no significant difference between CD4+ or CD8+ T cell density between airways or interstitium of the middle and lower lobe, respectively (**Fig. 3 C**). While there was a negative correlation between time since COVID-19 and airway T cell infiltrate, this trend was not statistically significant (p=0.1736).

### Assessment of interstitial fibrosis and quantification of macrophage subpopulations in PASC patients

Ultrastructural analysis (TEM) was performed on 11 of 14 PASC lung tissue samples with evidence of fibrosis in light microscopy and showed interstitial deposition of collagen fibrils in all cases (**Fig. 4 A** and **Tab. 2**). Immunofluorescence multiplex staining of lung tissue samples (n=38) for the quantification of macrophage subpopulations (**Fig. 4 B**) showed no significant differences between H-scores for pro-fibrotic macrophage phenotype (CD68/CD163/S100A9) according to clinical, imaging or serological data including diffusion capacity for carbon monoxide, oxygen pulse or number of CD4+/CD8+ T cells around airways or in alveolar septa (**Fig. 4 C** and **D**). The results from immunofluorescence multiplex staining are summarized in **Tab. 3**. The median proportion of CD163+ Macrophages among all Macrophages was 32%. Although this is higher than the median proportion of CD163+ Macrophages in a previously published control cohort (12%)[16], the difference was not statistically significant (p=0.2792). Analysis of cytokines in BAL fluid using legendplex immunoassay showed elevated levels of IL-1beta (p<0.05) and IL-8 compared to healthy controls, although the latter increase was not statistically significant (**Fig. 4 E**).

## DISCUSSION

In the present study, we report on the histopathological, serological, clinical and imaging characteristics in a consecutive cohort of 51 patients from the postCOVID outpatient clinic of the Bundeswehrkrankenhaus (army hospital) Ulm.

In comparison to most published data on the epidemiology of PASC [17], our patient cohort was younger (median age, 40 years) and the male sex was predominant (57%), both possibly due to the high proportion of active soldiers in the study and reflecting data from other studies involving military personnel [18].

Although all patients presented with dyspnea on exertion, pulmonary function tests revealed only limited pathological results. The most relevant finding were reduced MEF 50 and FEV1 (≤ 80%) in 42.9 and 16% of patients, in line with results from a previous study on athletes [19]. Airflow obstruction with a resulting reduction in MEF50/ FEV1 would be plausible concerning the observed proportion of patients with LAV>5% or >10% of lung area and the histopathologic finding of T cell-mediated bronchiolitis. It has been suggested that spiroergometry should be performed to assess cardiopulmonary function in PASC patients [20], but the median oxygen pulse was in the normal range. Since we had no pre-infection data, the actual impact of SARS-CoV-2 infection on individual cardiopulmonary function could not be assessed. Our data is in contrast to a study from Finland which did not find evidence of small airway inflammation in survivors of severe COVID-19 [21]. This difference might be due to the fact that we investigated a preselected cohort of patients with pulmonary symptoms after mild course of the disease who had not previously undergone anti-inflammatory treatment for severe COVID-19.

The observed correlation between oxygen pulse and time since infection in the complete cohort points towards normalization of oxygen pulse over time, possibly due to the parallel decrease in airway inflammation. In line with previous data [22], imaging in PASC patients showed GGOs and reticulations. There was no specific radiologic feature associated with clinical data, results from pulmonary function test or histopathological findings. In line with our finding of reduced MEF50/FEV1 in a subset of patients and our hypothesis of bronchiolitis-derived airflow obstruction, 35.3% of patients showed low attenuation volume (LAV) in >5% of lung area.

Both autoimmunity and persistence of virus/viral particles are discussed as possible contributors in the pathophysiology of PASC [11,13]. Using immunohistochemistry and RT-PCR, we did not detect SARS-CoV-2 or virus-derived spike or nucleocapsid proteins in lung tissue samples from all but one (2%) of PASC patients. In this patient the presence of SARS-CoV-2 could be confirmed by FISH in alveolar epithelium at 35 weeks after infection. Since this is a much longer time span than has previously been reported [23], it might be possible that the patient had a recent reinfection with SARS-CoV-2 despite negative RT-PCR from nasopharyngeal swab prior to inclusion into the present study.

We conclude that in the lung, persistence of virus/viral particles is not necessary for ongoing peribronchiolitis. This parallels the situation in acute COVID-19, where organization of diffuse alveolar damage/acute lung injury and long-term outcomes, such as pulmonary fibrosis occur after viral clearance and are therefore independent from the presence of virus [15]. With respect to autoantibodies, 33.3% of patients in the present study had detectable autoantibodies (ANA/ENA). Presence of autoantibodies was not associated with specific clinical characteristics, results from pulmonary function test or histopathological findings. Thus, they might reflect extrafollicular B cell activation upon contact with SARS-CoV-2 [24].

Histopathologic features of PASC were mostly discrete and non-specific, such as alveolar fibrin deposition and organizing pneumonia. When compared with pre-pandemic autopsy controls, there was significantly increased CD4+ (but not CD8+) T cell infiltrate around small airways (bronchiolitis), while interstitial CD4+/CD8+ T cell infiltrates were not significantly different. According to the classification proposed by Ryu [25], the inflammation pattern and imaging results reflected primary bronchiolitis rather than interstitial lung diseases with a prominent bronchiolar component. Previous viral or mycoplasmic infections are a frequent cause of primary bronchiolitis in children and adults and it has been shown that CD4+ T cells play a major role in obliterative bronchiolitis in chronic lung allograft dysfunction (CLAD) [26,27]. Bronchiolitis might also contribute to airflow obstruction that would explain the observed decrease in MEF50/FEV1 and increase of LAV areas in a subset of patients. Given 1) the established correlation between viral infections and persistent airway inflammation and 2) the absence of detectable SARS-CoV-2 in all but one patient, bronchiolitis in PASC seems to represent a rather unspecific post-infection reaction. Small peribronchiolar lymphoid follicles were present in about half of the patients. However, since interstitial inflammation was sparse, we would not see fulfilment of diagnostic criteria for IPAF/CTD-ILD that have previously been proposed [28,29]. It has to be noted, however, that no surgical lung biopsies were performed that would have allowed for recognition of cellular or fibrotic NSIP patterns in support of a diagnosis of IPAF/CTD-ILD. Finally, in a subset of patients, fibrotic remodeling could be detected by light microscopy and confirmed by TEM analysis; cytokine profiling of BAL fluid in an independent PASC cohort showed elevated levels of IF-1β. The proportion of CD163+ macrophage subpopulation in PASC lung tissue samples was higher than in a previously published control cohort, but this result was not statistically significant and the density of pro-fibrotic macrophages did not correlate with presence of fibrosis, clinical or imaging characteristics or the detection of autoantibodies. Still, we feel that pulmonary function should be monitored closely in PASC patients to allow for early detection and treatment of possible pulmonary fibrosis since it has been previously shown that SARS-CoV-2 has the ability to induce lung fibrosis [30,15].

There are several limitations to our study. First, we only included patients from a single center who had actively sought diagnostics and treatment of pulmonary symptoms after mild COVID-19, so it is impossible to draw any conclusions about the frequency and severity of pulmonary PASC in the general population. Second, the significance of the findings for an individual patient might be limited due the high number of physically active soldiers in our cohort who might have less tolerance for even slight impairment of lung function. On the other hand, the absence of relevant pulmonary comorbidities in this particular group and the fact that infections occurred before the widespread roll-out of vaccines offered a unique opportunity to study the natural course of lung pathology after SARS-CoV-2 infection. Third, since most patients were infected during the alpha/delta waves, it is unclear whether the findings hold true for infections with omicron or other subtypes of SARS-CoV-2. Finally, inclusion of lung tissue samples from patients after non-COVID viral infections as controls would have allowed us to investigate to which extent post-infection bronchiolitis/fibrosis is COVID-specific, however these patients do not usually undergo biopsy. We will follow-up the present cohort to investigate whether bronchiolitis and autoantibodies persist and to monitor whether there is clinical evidence of progressive fibrosis especially in the subset of patients who had first evidence of fibrosis in the baseline biopsy.

## Data Availability

All data produced in the present study are available upon reasonable request to the authors

## SUMMARY AND INTERPRETATION

We report here on a consecutive cohort of 51 unvaccinated patients who presented with pulmonary PASC, mostly in form of dyspnea on exertion. Only a minority of these patients had significantly impaired lung function, however a significant subset had impaired MEF50/FEV1 and enhanced LAV, in line with airflow obstruction, possibly due to T cell-mediated bronchiolitis. Residual SARS-CoV-2 was not detectable in lung tissue of all but one PASC patient, but a subset of patients showed evidence of fibrotic remodeling of the lung. We think that obstructive lung disease due to post-infectious bronchiolitis without viral persistence is the main cause of pulmonary PASC after mild course of the disease. While in most cases pulmonary symptoms might be self-limiting and wane over time, there is evidence of early fibrotic remodeling in a subset of PASC patients.

## ACKNOWLEDGEMENTS

The authors would like to thank all patients for their consent to the use of data and images in the present study.

## AUTHOR CONTRIBUTIONS

Study concept: DG, WB, PB and KS. Data collection: DG, CH, AC, AL, NM, WB, FZ, FK, SD, SvS, RB, PB, KS. Sample collection: DG, NM, SvS, KS. Initial draft of manuscript: KS. Critical revision and approval of final version: all authors.

